# Peripheral blood *GATA2* expression impacts *RNF213* mutation penetrance and clinical severity in moyamoya disease

**DOI:** 10.1101/2024.06.22.24306750

**Authors:** Yohei Mineharu, Takahiko Kamata, Mei Tomoto, Noriaki Sato, Yoshinori Tamada, Takeshi Funaki, Yuki Oichi, Kouji H Harada, Akio Koizumi, Tetsuaki Kimura, Ituro Inoue, Yasushi Okuno, Susumu Miyamoto, Yoshiki Arakawa

## Abstract

**Background:** The p.R4810K founder mutation in the *RNF213* gene confers susceptibility to moyamoya disease (MMD) and non-MMD intracranial artery disease. However, penetrance is incomplete, and the underlying molecular mechanism remains unknown.

**Methods and Results:** Transcriptome analysis of peripheral blood was conducted with 9 MMD patients and 5 unaffected mutation carriers from 4 familial MMD pedigrees. Bayesian network analysis identified upregulated gene modules associated with lipid metabolism and leukocyte development (including *GATA2* and *SLC45A3*), and EGFR signaling (*UBTD1*). It also identified downregulated gene modules related to mitochondrial ribosomal proteins (*RPS3A* and *RPL26*), and cytotoxic T cell immunity (*GZMA* and *TRGC1*). The *GATA2* network was replicated through WGCNA analysis and further examined in a case-control study, comprising 43 MMD patients, 16 non-MMD patients, 19 unaffected carriers, and 35 healthy controls. *GATA2* exhibited a significant linear correlation with *SLC45A3* and was significantly higher in MMD patients compared to age- and sex-matched unaffected carriers or wild-type controls. Among patients with the p.R4810K mutation, higher *GATA2* expression was associated with an earlier age of onset, bilateral involvement, and symptomatic disease onset.

**Conclusions:** Peripheral blood *GATA2* expression was associated with increased penetrance of the *RNF213* mutation and more severe clinical manifestations in MMD.

## Introduction

Moyamoya disease (MMD) is a progressive cerebrovascular condition characterized by steno- occlusive lesions at the terminal portion of the internal carotid arteries,^1,2^ with the development of multiple collateral networks, including basal moyamoya vessels and periventricular anastomoses.^3^ The age distribution for MMD shows two peaks: those under 10 years old, who are more likely to experience ischemic stroke, and those in their 30s and 40s, who are more prone to hemorrhagic stroke, although there is some variation across different ethnicities.^4,5^ While the exact pathophysiology of MMD is not yet fully understood, various related molecules have been gradually identified.^2,6^ The *RNF213* gene is a major susceptibility gene for MMD, with the founder mutation p.R4810K occurring in approximately 90% of Japanese patients, 70% of Korean patients, and 40% of Chinese patients.^7,8^ By contrast, other minor mutations are found in only 10% of non-East Asian patients. The high prevalence of MMD in East Asia is presumed to be associated with the high frequency of the founder mutation. Despite the fact that the p.R4810K founder mutation in the *RNF213* gene increases the risk of MMD by 300 times, its penetrance is incomplete. Given that the p.R4810K mutation is found in 1-2% of the general Japanese population, the penetrance of the heterozygous mutation is estimated to be around 0.5- 1.0%.^9^ This has prompted a search for factors that could modify penetrance, but none have been definitively identified. Furthermore, the p.R4810K mutation is associated not only with MMD but also with non-moyamoya intracranial large artery disease (ICAD).^10,11^ Identifying modifier genes for MMD could provide insights into the differences between MMD and ICAD.

Recent studies have indicated that the immune system is involved in the pathogenesis of MMD,^12–18^ and *RNF213* plays a role in regulating immune responses to intracellular pathogens.^19,20^ For example, *RNF213* has been reported to ubiquitinate non-protein substrates such as lipopolysaccharide (LPS) on Salmonella.^21^ *RNF213* is highly expressed in human immune cells, and a mouse model showed that *RNF213* mutations affect dendritic cell function.^22^ The association of certain microbiota, such as *Ruminococcus gnavus* (*R. gnavus*) and *Campylobacter*, with MMD also suggests that systemic factors may play a role in the pathogenesis of MMD.^23–25^ Based on this, we hypothesized that transcriptional changes in peripheral blood might influence the penetrance of the p.R4810K mutation. In the present study, we analyzed the transcriptomic differences between patients with MMD and unaffected mutation carriers from large families with familial MMD. Possible modifier genes for the *RNF213* mutation were identified using Bayesian network analysis, a machine learning technique, and the association between the gene expression and MMD phenotypes was confirmed in additional non-familial cases.

## PATIENTS and METHODS

### Study population and sample collection

The study was conducted in accordance with the Declaration of Helsinki and Japanese Ethical Guidelines for Medical and Health Research Involving Human Subjects. Approval for the study was obtained from the ethics committee of the Institutional Review Board (G0737 and R2088; Kyoto University, Kyoto, Japan), and all participants provided written informed consent. If the participants were minors, informed consent was given by their parents and/or regal guardians along with informed assent. Among the 19 families analyzed in our previous linkage study,^26^ four families with incomplete penetrance of the p.R4810K mutation were selected for transcriptome analysis (Figure 1). This group comprised 9 patients with MMD (#3, 4, 6, 7, 10, 12, 13, 15, 16) and five carriers (#2, 5, 9, 11, 14). Patient #2 later developed MMD, and patient #5 had middle cerebral artery stenosis, but at the time of participation, neither met the criteria for MMD, so they were classified as unaffected. Patients #1 and #8 provided blood samples, but they were excluded because the RNA extraction from these samples did not meet the quality standards. Gene expression of candidate genes identified by the transcriptome analysis was measured in a prospectively recruited replication cohort consisting of 43 patients with MMD, 16 patients with ICAD, 19 unaffected mutation carriers, and 35 controls with other comorbidities but no history of cerebrovascular diseases. For age- and sex-matched analysis, participants were selected such that the age range was within 10 years and gender was matched. Peripheral blood samples (4-9ml) were collected from the participants, and samples for transcriptome analysis were collected into PAXgene Blood RNA tubes (BD Bioscience, #762165, Franklin Lakes, NJ, USA).

**Figure 1.**
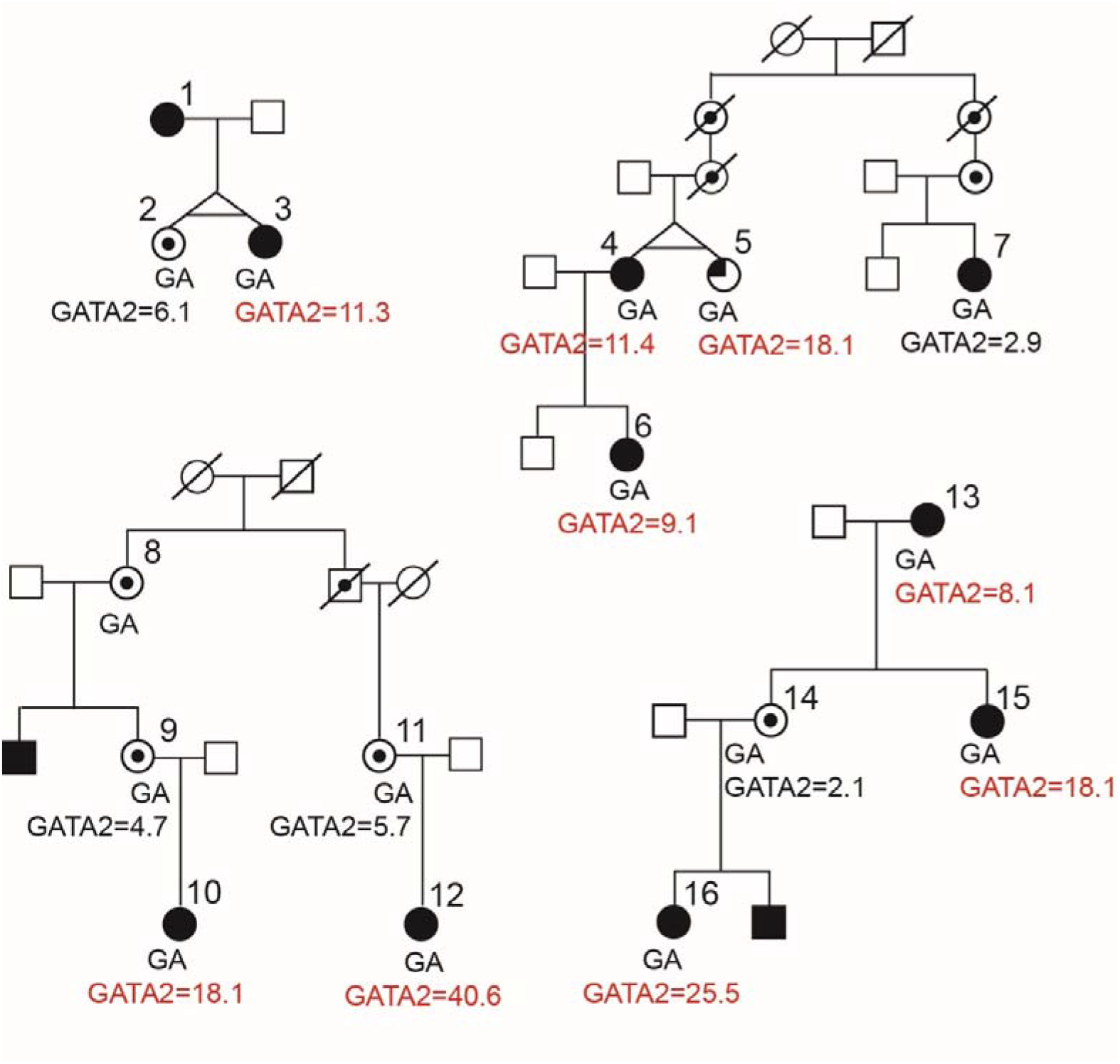
Pedigree Chart of Familial MMD with Incomplete Penetrance. Peripheral blood was collected from family members with familial moyamoya disease (MMD). All MMD patients had a relative *GATA2* expression level above 8.06 except for one patient (#7). Unaffected mutation carriers (#2, 9, 11, 14) had levels below 8.06. Although we treated patient #5 who had middle cerebral artery stenosis as unaffected, she showed *GATA2* expression level above 8.06. RNA samples from individuals #1 and #8 were not obtained. Filled symbols represent patients with MMD; dotted symbols represent unaffected carriers of the *RNF213* p.R4810K mutation; the quarter-filled symbol represents a case with unilateral middle cerebral artery stenosis. Below the symbols, the p.R4810K mutation genotype (heterozygote, GA) and the relative *GATA2* expression values.

### Transcriptome analysis

Total RNA was collected from peripheral blood using the PAXgene Blood RNA kit (BD Bioscience #763134). RNA was extracted according to the manufacturer’s instructions and kept frozen until use. The NEBNext® Poly(A) mRNA Magnetic Isolation Module was used to isolate mRNA from the total RNA. The library was created using the NEBNext Ultra II RNA Library Prep Kit for Illumina. RNA-Seq was performed on the NovaSeq 6000 with 100bp single-end reads (NovaSeq6000 SP Reagent Kit v1.5 (100 Cycles)). Depending on the quantity of cells homogenized in RLT buffer (Qiagen, #79216, Hilden, Germany), total RNA was isolated using either the RNeasy Mini Kit (Qiagen, #74106) or the RNeasy Micro Kit (Qiagen, #74004), with an on-column DNase digestion (Qiagen, #79254), following the manufacturer’s instructions. The quality and quantity of the total RNA were assessed using the Agilent 2100 Bioanalyzer with the "Agilent RNA 6000 Nano" kit (G2938-90034, Agilent Technologies, Santa Clara, CA, USA) and the "Agilent RNA 6000 Pico" kit (G2938-90046, Agilent Technologies). Total RNA samples with an RIN > 7 were used to prepare the TruSeq stranded mRNA libraries according to the manufacturer’s instructions (Document # 1000000040498 v00, Illumina). The stranded mRNA libraries were analyzed on an Agilent 2100 Bioanalyzer using the "Agilent DNA 7500" kit (G2938-90024, Agilent Technologies), diluted to 10 nM, and pooled equimolar into multiplex sequencing pools for sequencing on the HiSeq 2500 and NovaSeq 6000 Illumina sequencing platforms. Paired-end sequencing on the NovaSeq 6000 used 54 cycles for Read 1, 19 cycles for Read i7, 10 cycles for Read i5, and 54 cycles for Read 2, with the NovaSeq6000 SP Reagent Kit v1.5 (100 cycles) (Illumina, #20028401, San Diego, CA, USA). RNA libraries were prepared for sequencing using standard Illumina protocols. The average cluster number was 69,958,858. Quality control was performed using Trimmomatic with the following parameters: LEADING:20, TRAILING:20, SLIDINGWINDOW:4:15, MINLEN:36. Adapter trimming was performed with ILLUMINACLIP:adapters.fa:2:10:10.^27^ Reads were mapped to the genome with GRCh38 using STAR.^28^ Read counts were calculated by featureCounts,^29^ and normalized by the trimmed mean of M-values (TMM) using edgeR.^30^

### Bayesian network analysis

INGOR.0.14.0 (A newer version of SiGN-BN) was used for Bayesian network estimation.^31–34^ The super-computing resource was provided by the Human Genome Center, the Institute of Medical Science, the University of Tokyo (http://sc.hgc.jp/shirokane.html). The Basal network was estimated by a Bayesian network (BN) using the B-spline nonparametric regression model, a graphical model that represents conditional independence among variables as a directed acyclic graph, where directed edges can be thought of as representing causal relationships among nodes. BN searches for conditional independence among variables to estimate causal relationships among a large number of variables. That is, given multiple factors (variables) *X*_1_, *X*_2_, …*X_p_* are given, their simultaneous probability *Pr*(*X*_1_, *X*_2_, …*X_p_)* is expressed by the following equation.

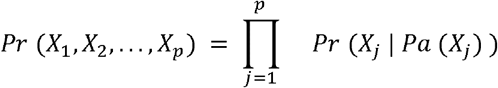

where *p* is the number of variables and *PaX_j_)* is the parent of *X_j_*

For a nonparametric BN with continuous values as input, the simultaneous probabilities are expressed by decomposing the probability density function as follows:

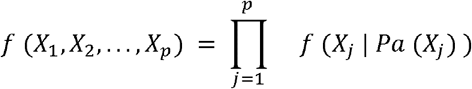

The network structure that matches the data well, i.e., has a high posterior probability, was estimated as a Basal Network.

The Neighbor Node Sampling and Repeat (NNSR) algorithm was employed for BN estimation. The NNSR algorithm is a method that can estimate networks from large data sets, which is not possible with other algorithms, by iteratively estimating sub-networks of selected genes by sampling neighboring nodes in parallel.^32^ The cutoff value was set at 0.05. The number of subnetwork estimation iterations (denoted as T) was set to T = 100,000, and network estimation was performed three times under identical conditions. Edge agreement was calculated for each pair of estimated networks, and it was confirmed that, on average, more than 95% of the network structures were consistent. This indicates that the estimated network has a sufficiently stable structure. The final basal network structure was obtained by reducing the number of nodes and edges by ignoring invalid scores such as infinity and NaN.

Extraction of subnetworks using Edge Contribution value (ECv) was done as follows.

In the nonparametric regression model of the estimated network, the parent-child relationship between variables is expressed by the following relational equation

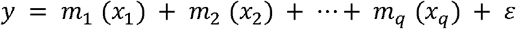

where *m_q_* (*x_q_*) is the value that quantifies the usage of the edge for each sample of edge *x_q→_y*. This value is called ECv and the ECv for *x_k→_y* w.r.t. sample u :is defined by the following equation

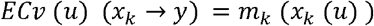

*x_k_*(*u*) the value of the *k*-th parent of *Y* in sample *u* .

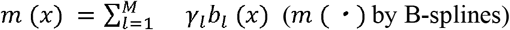

between different conditions. At edge *x_k_→y*, ΔECv, the difference between ECvs in sample In a previous study, ECvs were compared to identify differences in the networks of two samples groups s and *t* between different conditions, is calculated by the following formula:

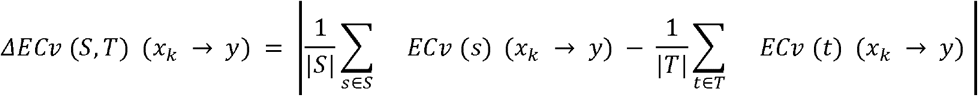

By extracting edges with ΔECv above a threshold value and the nodes connected to them, we can extract subnetworks that move characteristically under certain conditions. In past analyses, a threshold value of 1.0 for ΔECv has been used.^34^ Visualization of the estimated network was performed using Cytoscape.^35^

### WGCNA analysis

We applied Weighted Gene Co-Expression Network Analysis (WGCNA) to construct gene co-expression networks and identify consensus modules and hub genes that play significant roles in enhancing the penetrance of the p.R4810K mutation, using RNA-Seq datasets. The WGCNA process begins by creating an adjacency matrix, using a selected soft-thresholding power to maintain an approximate scale-free topology, and constructing a topological overlap matrix (TOM) to reduce noise and minimize spurious correlations. The analysis used preprocessed and filtered data as input. To determine the optimal soft-thresholding power, we tested values ranging from 1 to 30. The scale-free topology fit index (SFTI) and mean connectivity were then calculated, with a chosen power at which the SFTI first exceeded 0.8 across all datasets. This determined power was then used to calculate the TOM. We selected a "signed" TOM type, a "signed hybrid" network, and used the biweight midcorrelation method for correlation calculations. The TOM was scaled, and a consensus TOM was defined by taking the component-wise minimum across all datasets. By subtracting this consensus TOM from one, we created a new matrix and performed hierarchical clustering on it. The resulting dendrogram was pruned using the cutreeDynamic function, with a minimum cluster size of 35 and a deepSplit parameter of 2. Following this, we calculated eigengenes, which represent the first principal components of the expression values for each cluster, and derived the correlation matrix for these eigengenes. This correlation matrix was then subtracted from one, followed by a second hierarchical clustering to merge closely related modules, with a maximum dissimilarity parameter of 0.5. Finally, these modules were used to perform linear regression analyses, with the module eigengenes as the dependent variable and disease status as the independent variable, to identify which eigengenes differed between disease states in each dataset. The relationships among the module eigengenes were visualized using the R library pvclust.

### RT-qPCR

The extracted total RNA PAXgene Blood RNA kit was reverse transcribed into complementary DNA (cDNA) using Superscript III (Invitrogen, Waltham, MA, USA). The expression of target genes was normalized using *RPS18* as the reference gene. Quantitative PCR (qPCR) was performed with KOD SYBR qPCR Mix (TOYOBO, Osaka, Japan) on a StepOnePlus system (Thermo Fisher Scientific), following the Delta Delta Ct method. The PCR cycling conditions adhered to the recommendations provided by TOYOBO. The forward and reverse primer sequences used for amplification are summarized in Supplemental Table 1. Relative expression was calculated using the value from the patient #CVD004).

### Genotyping

DNA was extracted from peripheral blood samples using the QIAamp DNA Blood Mini Kit (Qiagen, Germantown, Maryland, USA), following the manufacturer’s instructions. The quality and concentration of the extracted DNA were assessed using the Infinite M200 PRO (TECAN, Kanagawa, Japan) or Qubit Flex Fluorometer (Thermo Fisher Scientific, Waltham, MA, USA). The extracted DNA was stored at -30 °C until further analysis. Genotyping for the p.R4810K mutation was performed for all participants using a TaqMan probe (Custom TaqMan SNP

Genotyping Assays; Applied Biosystems, Foster City, CA, USA) and a 7300/7500 Real-Time PCR System (Applied Biosystems), as previously described and in accordance with the manufacturer’s instructions.^36^

### 16S rRNA Metagenomic Analysis

Fecal samples were obtained from 48 individuals (26 MMD patients, 5 family members, 9 ICAD patients, and 16 controls). The samples were collected in stool sample containers (FS- 0014, TechnoSuruga Laboratory Co., Ltd., Shizuoka, Japan) and stored at -80 °C. Details regarding the 16S rRNA metagenomic analysis have been described elsewhere.^23^ Briefly, DNA was extracted from the fecal samples using the NucleoSpin DNA Stool Kit (Takara Bio Inc., Shiga, Japan), following the manufacturer’s instructions. The DNA was suspended in a 30 µl extraction buffer and stored immediately at -80 °C. The DNA was amplified using a two-step tailed PCR targeting the V3-V4 regions of bacterial 16S rRNA. Each PCR reaction was carried out with Ex Taq HS (Takara Bio Co., Shiga, Japan). V3-V4 PCR products were purified using AMPure magnetic beads (Beckman Coulter, United States) and quantified using a Synergy H1 (BioTek, United States) and the QuantiFluor dsDNA System (Promega, United States).

Sequencing was conducted with the MiSeq Reagent Kit v3 (Illumina, United States) under the 2 x 300 bp configuration. Using the fastx_barcode_splitter tool from the FASTX Toolkit (ver. 0.0.0.14), we extracted only those read sequences that exactly matched the primer sequence used at the beginning of the read. Sequences with a quality value below 20 were removed using Sickle (ver. 1.33), and those shorter than 130 bases were discarded. Paired-end reads were joined using FLASH (ver. 1.2.11). After removing chimeric and noisy sequences using the dada2 plugin in Qiime2 (ver. 2021.8), representative sequences and amplicon sequence variant tables were generated. Taxonomic classification was performed using the feature-classifier plugin, and the alignment and phylogeny plugins were used to create phylogenetic trees. We calculated relative abundances from the raw count matrix.

### Statistical Analysis

Categorical variables were analyzed using either the chi-squared test or Fisher’s exact test, depending on the context. Continuous variables were compared using the Student’s t-test with Welch’s correction. The Cochran–Armitage test was applied to detect linear trends in categorical variables based on *GATA2* expression levels, while Welch’s Analysis of Variance (ANOVA) was used for continuous variables, with *GATA2* expression levels categorized into low (0-8), medium (8-16), and high (>16). Receiver operating characteristic (ROC) analysis was used to determine the optimal cutoff value for *GATA2* expression in distinguishing patients from unaffected mutation carriers. The optimal cutoff on the ROC curve was identified as the point that maximizes the vertical distance from the diagonal line (where Sensitivity = 1 - Specificity). Pearson’s correlation coefficient (r) was used to assess linear correlations between continuous variables. A *p*-value of less than 0.05 was considered statistically significant. All statistical analyses were conducted using R version 4.0.5 and GraphPad Prism version 9.3.1.

## RESULTS

### Bayesian Network Analysis identified gene networks that affect the p.R4810K penetrance

RNA-Seq was conducted with 10 patients with MMD and 5 unaffected mutation carriers (Figure 1). The clinical characteristics of the study population are shown in Table 1. The unaffected carriers were significantly younger than the patients with MMD. Differentially expressed genes (DEGs) between patients and carriers were analyzed. After adjustment for multiple comparisons, only seven genes met the significance threshold, and the heatmap did not reveal any specific pattern distinguishing MMD patients from unaffected carriers. We considered that the small sample size might have resulted in insufficient statistical power, limiting the identification of disease-related gene expression. Since the expression of certain genes is closely interrelated, grouping these genes might increase statistical power.

**Table 1.**

|  | Age- and sex-matched population |  |  |  |  | Total population |  |  |  |  |
| --- | --- | --- | --- | --- | --- | --- | --- | --- | --- | --- |
|  | Carrier<br>(mutant) | MMD | *P* | Control<br>(wildtype) | *P* | Control | Family | ICAD | MMD | *P* |
| Number, n | 10 | 10 |  | 10 |  | 35 | 19 | 16 | 43 |  |
| Age, median (IQR) | 47.0 (22.0) | 53.0 (23.5) | 0.35 | 45.5 (17.5) | 0.21 | 56 (33.0) | 37 (28.0) | 51 (25.25) | 37 (30.5) | < 0.01 |
| Female, n (%) | 7 (70.0) | 7 (70.0) | 1 | 7 (70.0) | 1 | 19 (54.3) | 10 (52.6) | 10 (62.5) | 35 (81.4) | 0.035 |
| Hypertension, n (%) | 2 (20.0) | 3 (30.0) | 1 | 4 (40.0) | 0.63 | 9 (25.7) | 3 (15.8) | 5 (31.3) | 7 (16.3) | 0.5 |
| Diabetes, n (%) | 0 (0) | 0 (0) | 1 | 1 (10.0) | 1 | 2 (5.7) | 0 (0) | 1 (6.3) | 4 (9.3) | 0.62 |
| Dyslipidemia, n (%) | 0 (0) | 1 (10.0) |  | 1 (10.0) | 1 | 3 (8.6) | 0 (0) | 8 (50.0) | 5 (11.6) | < 0.01 |
| Drinking, n (%) | 1 (10.0) | 1 (10.0) | 1 | 1 (10.0) | 1 | 8 (22.9) | 2 (10.5) | 2 (12.5) | 5 (11.6) | 0.55 |
| Smoking, n (%) | 1 (10.0) | 3 (30.0) | 0.58 | 3 (30.0) | 0.58 | 11 (31.4) | 2 (10.5) | 6 (37.5) | 7 (16.3) | 0.11 |
| Family history, n (%) | 8 (80.0) | 7 (70.0) | 1 | 0 (0) | <0.01 | 0 (0) | 19 (100) | 4 (25.0) | 25 (58.1) | < 0.01 |
| *RNF213* p.R4810K, n (%) | 10 (100) | 10 (100) | 1 | 0 (0) | <0.01 | 2 (5.7) | 10 (52.6) | 7 (43.8) | 40 (93.0) | < 0.01 |

MMD represents moyamoya disease; ICAD, intracranial large artery disease. Family history indicates family history of MMD.

Next, we used Bayesian network analysis to identify gene networks associated with the increased penetrance of the p.R4810K mutation. The complete network was successfully visualized using the INGOR program (Figure 2a). The Delta Edge Contribution value (ΔECv) represents the difference in ECv between patients and unaffected mutation carriers, allowing for the identification of patient-specific networks that differ from those of unaffected carriers. The top 1% of the network by ΔECv was depicted (Figure 2b). Networks with ΔECv >1.0 were considered significant, leading to the identification of four key networks (Figures 2c-f). These included a downregulated network of mitochondrial ribosomal proteins (*RPS3A*, *RPL23*, and *RPL26*) and a downregulated T cell immunity network (*GZMA*, *APOBEC3H*, and *TRGC1*).

**Figure 2.**
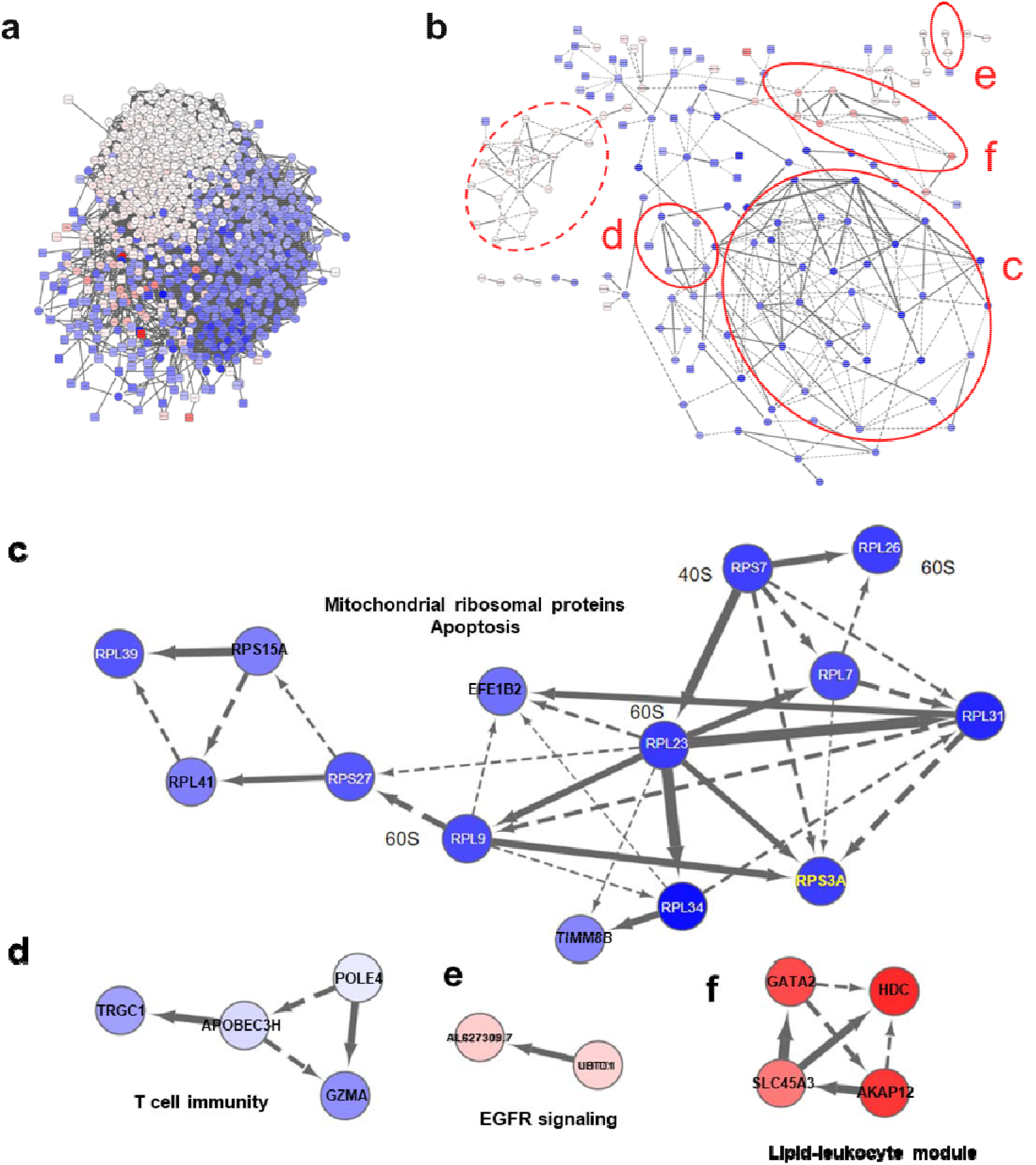
Bayesian Network Analysis Identified 4 Major Gene Networks. A Bayesian network analysis using INGOR was performed to identify patient-specific gene networks, and the complete network was successfully visualized (a). ΔECv, indicating the difference in Edge Contribution value (ECv) between patients and unaffected mutation carriers, was calculated, and the top 1% of the network was depicted (b). Among them, networks with ΔECv >1.0 were considered significant, leading to the identification of four major networks (solid circle, c-f). These included a network of mitochondrial ribosomal proteins such as *RPS3A*, *RPL23*, and *RPL26* (c), a T cell immunity network featuring *GZMA*, *APOBEC3H*, and *TRGC1* (d), an EGFR signaling network with *UBTD1* (e), and a lipid-leukocyte module comprising *GATA2*, *SLC45A3*, *HDC*, and *AKAP12* (f). Genes highlighted in red are upregulated, while those in blue are downregulated. Although it did not reach the significance threshold, a subnetwork encompassing IL-1β and SOCS3 was detected (dotted circle, details in Supplementary Figure 2).

Additionally, two upregulated networks were identified: an EGFR signaling network (*UBTD1*), and a lipid-leukocyte module (*GATA2*, *SLC45A3*, *HDC*, and *AKAP12*) which is involved in both lipid metabolism and leukocyte development. Although it did not reach the significance threshold, another interesting network emerged (Figure 2b, dotted circle; details in Supplementary Figure 1), which included the upregulation of *IL-1*β and *SOCS3*.

### Association of GATA2 with MMD and increased penetrance of the RNF213 mutation

The significant association of the lipid-leukocyte module with increased penetrance of the p.R4810K mutation was confirmed through WGCNA analysis (Figure 3a). RT-qPCR analysis also showed a strong correlation between *GATA2* and *SLC45A3* expression levels (Figure 3b). We then explored the association of *GATA2* expression with MMD, including patients with intracranial large artery disease (ICAD), a type of RNF213-related vasculopathy. *GATA2* expression was measured using RT-qPCR, and the levels were compared among MMD patients with the mutation, age- and sex-matched unaffected mutation carriers, and wild-type controls without cerebrovascular diseases. As shown in Figure 3c, *GATA2* expression was similar between mutation carriers and wild-type controls (*p* = 0.896), but significantly higher in patients with MMD compared to mutation carriers (*p* = 0.0285). ROC analysis revealed an area under the curve (AUC) of 0.74, with an optimal cutoff for relative *GATA2* expression of 8.06 to distinguish MMD patients from unaffected mutation carriers (Figure 3d). This cutoff value was applicable to the original familial populations (Figure 1), where all individuals with *GATA2* expression greater than 8.06 had MMD except for one patient who had middle cerebral artery stenosis (#5), and all individuals with *GATA2* expression less than 8.06 did not have MMD except for one patient with MMD (#7).

**Figure 3.**
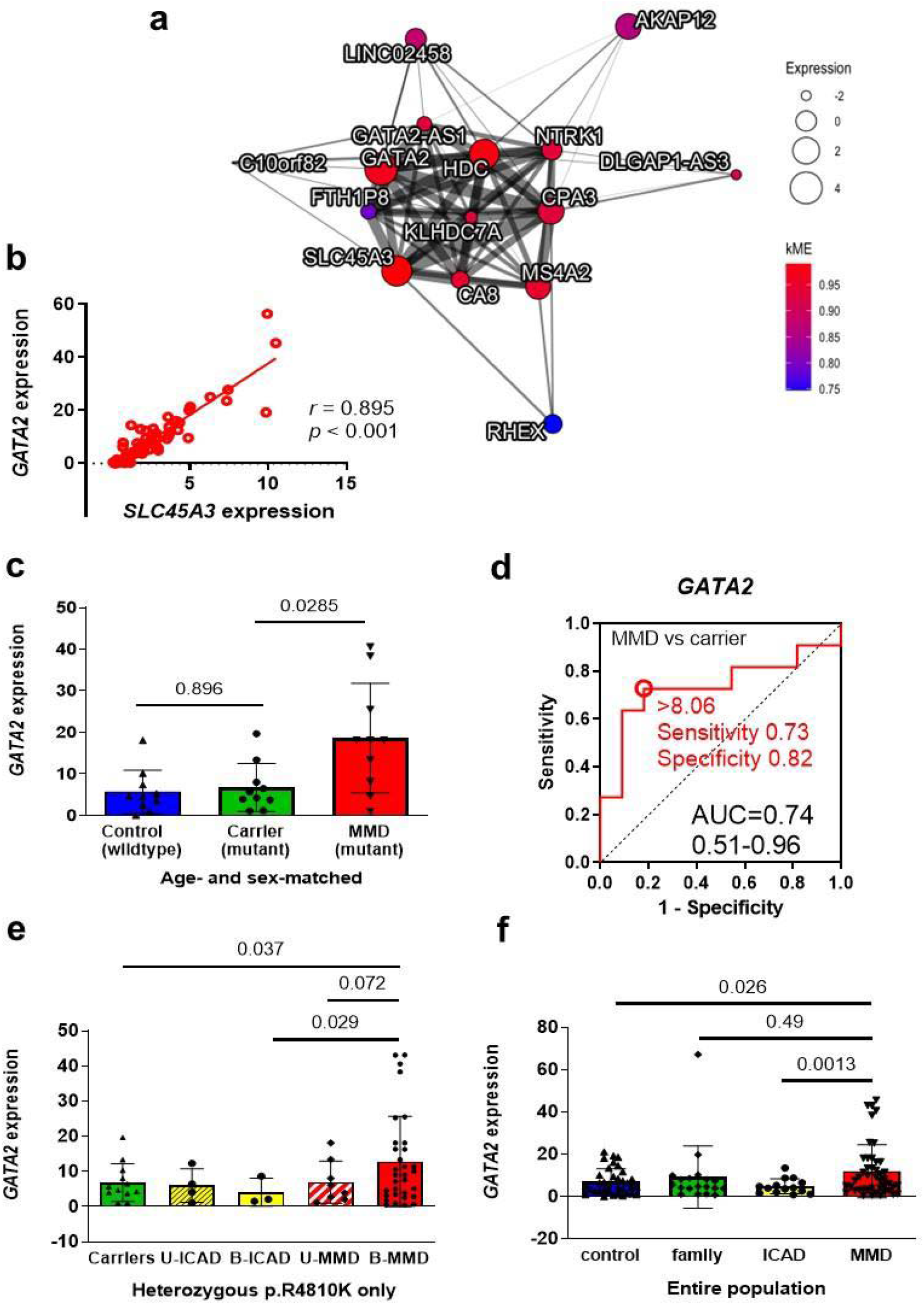
Significance of *GATA2* Gene Expression to Differentiate MMD Patients and Unaffected *RNF213* Mutation Carriers. The Weighted Gene Co-Expression Network Analysis (WGCNA) replicated the results of the Bayesian network analysis using INGOR, identifying the lipid lymphocyte (LL) module, which includes *GATA2* and *SLC45A3*, as a significant network (a). A strong correlation between *GATA2* and *SLC45A3* expression was observed, with r = 0.895 (p < 0.001) (b). Peripheral *GATA2* expression was compared among 10 patients with moyamoya disease (MMD), 10 age- and sex-matched controls, and 10 unaffected mutation carriers. *GATA2* expression was significantly higher in patients with MMD compared to controls (*p* = 0.027) and unaffected carriers (*p* = 0.045; Dunnett’s multiple comparison test) (c). The ability of *GATA2* expression to distinguish patients with MMD from unaffected carriers was assessed using Receiver Operating Characteristic (ROC) analysis. The area under the curve was 0.74, with the optimal cutoff set at 8.06 (d). *GATA2* expression was compared among individuals with the heterozygous p.R4810K mutation, revealing that patients with bilateral MMD (B-MMD) had higher *GATA2* expression than controls or patients with bilateral non-moyamoya intracranial arterial disease (B-ICAD) (e). *GATA2* expression was then compared across the entire study population, including individuals with homozygous and wild-type *RNF213* alleles (f).

We further examined *GATA2* expression among individuals with the heterozygous p.R4810K mutation. *GATA2* expression in patients with bilateral MMD was higher than in unaffected carriers, and it was also higher than in those with bilateral ICAD (Figure 3e). This significance persisted when the entire study population was considered, with patients with MMD showing higher *GATA2* expression than both controls and patients with ICAD (Figure 3f). The difference between MMD patients and their family members might be due to the higher expression level in family members with the wild-type allele, suggesting that elevated *GATA2* expression appears to increase the risk of MMD in the presence of the p.R4810K mutation.

### Clinical Significance of *GATA2* Expression in RNF213-Related Vasculopathy

We then assessed the clinical significance of *GATA2* expression in RNF213-related vasculopathy, including MMD and ICAD. When we divided the patients into three groups based on relative *GATA2* expression (0 to 8, 8 to 16, and ≥16), higher expression of *GATA2* was significantly associated with younger age at onset (*p* = 0.0353, Figure 4a). To determine whether elevated *GATA2* expression in early-onset patients might be influenced by the age at blood collection rather than its association with the disease, we examined *GATA2* expression levels in a control group. The results indicated that *GATA2* expression was higher in older individuals (Figure 4b), suggesting that higher *GATA2* levels in early-onset cases are linked to the disease and not confounded by the age at blood collection. We also investigated the relationship between *GATA2* expression and disease duration (the time between disease onset and blood collection) and found no significant correlation (Figure 4c), confirming that variability in this interval does not strongly affect *GATA2* expression. When stratifying *GATA2* expression by gender and age (children and adults), expression levels were higher in boys during childhood and slightly higher in women during adulthood, with a significant interaction effect. In an analysis limited to patients, the same trend was observed, although the difference was not statistically significant (Supplementary Figure 2). Higher *GATA2* expression was also associated with a higher proportion of bilateral involvement and symptomatic onset (Figures 4d, e), although the linear trend was not statistically significant. The association between *GATA2* expression and clinical features became less apparent when patients with ICAD were excluded (Supplemental Figure 3).

**Figure 4.**
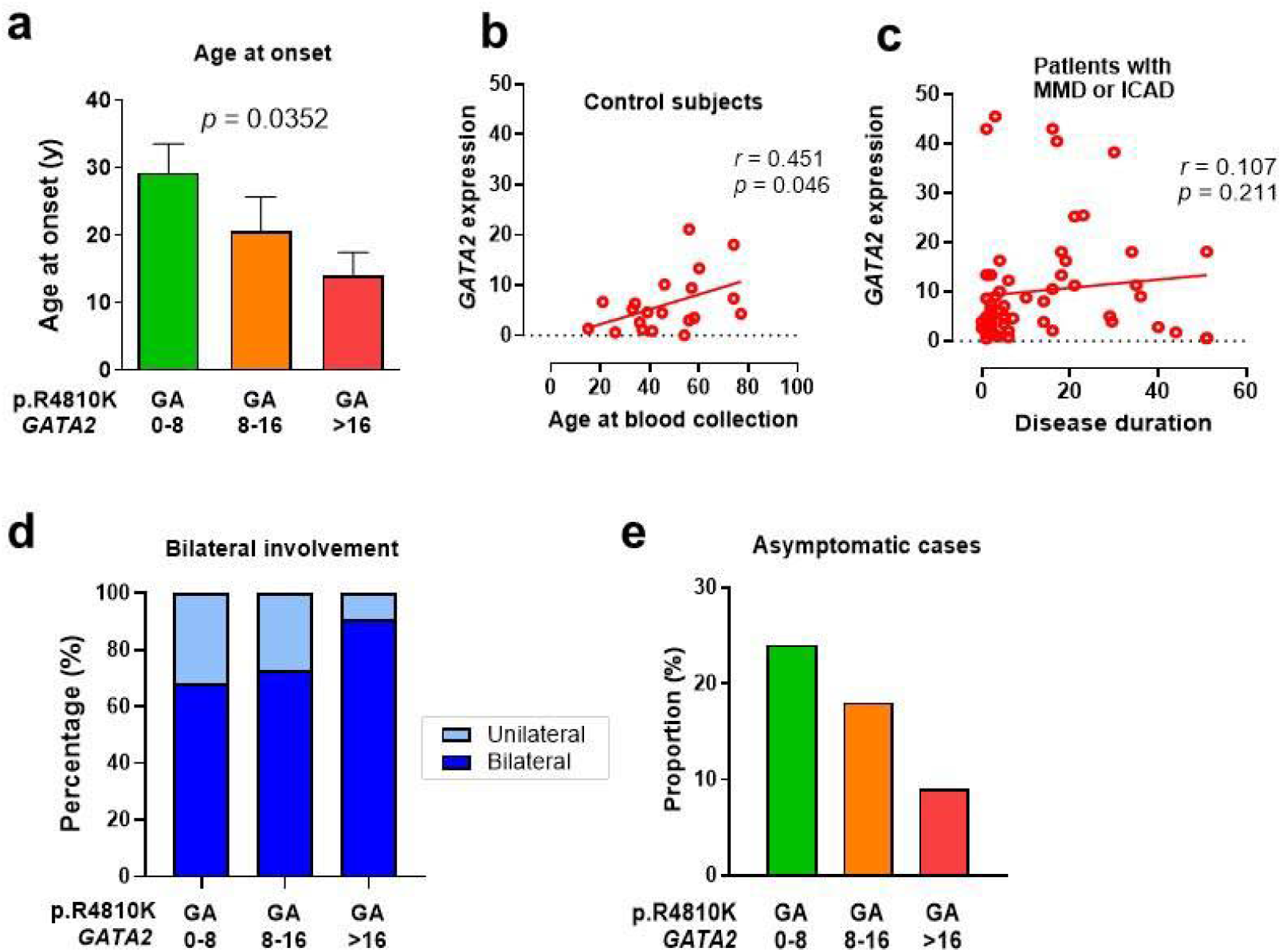
Association of Peripheral Blood *GATA2* Expression Levels and Disease Severity in Patients with MMD or ICAD with a Heterozygous p.R4810K Mutation in the *RNF213* Gene. *GATA2* expression was divided into three groups (n = 25 for *GATA2* 0-8; n = 11 for *GATA2* 8- 16; n = 11 for *GATA2* > 16). A higher *GATA2* expression was associated with a younger age at onset, according to Welch’s ANOVA test (a). In controls, *GATA2* expression decreased with age, suggesting that the inverse correlation between *GATA2* expression and age at onset is not influenced by the age at which the blood samples were collected (b). *GATA2* expression did not correlate with disease duration, defined as the time between age at onset and age at blood collection (c). A higher *GATA2* expression was linked to a trend toward a higher proportion of bilateral involvement (68.0%, 72.7%, and 90.9%), although the difference was not statistically significant (d). Similarly, a higher *GATA2* expression showed a trend toward a higher proportion of symptomatic cases, but this trend was also not statistically significant (e).

### Relationship between peripheral *GATA2* levels and *Ruminococcus gnavus* in the gut

In our previous studies, we reported that the relative abundance of *R. gnavus* is associated with MMD.^23^ It has also been reported that *R. gnavus* promotes the secretion of type 2 cytokines, IL- 5 and IL-13, from CD4+ T cells.^37^ Given that *GATA2* is involved in type 2 inflammation, which encompasses eosinophils and mast cells, we hypothesized a potential connection between *R. gnavus* and *GATA2*. Upon analyzing the correlation between the relative abundance of *R. gnavus* and *GATA2* expression, a weak but significant correlation was observed (Figure 5a). However, when the analysis was restricted to patients only, the statistical significance disappeared (Figure 5b).

**Figure 5.**
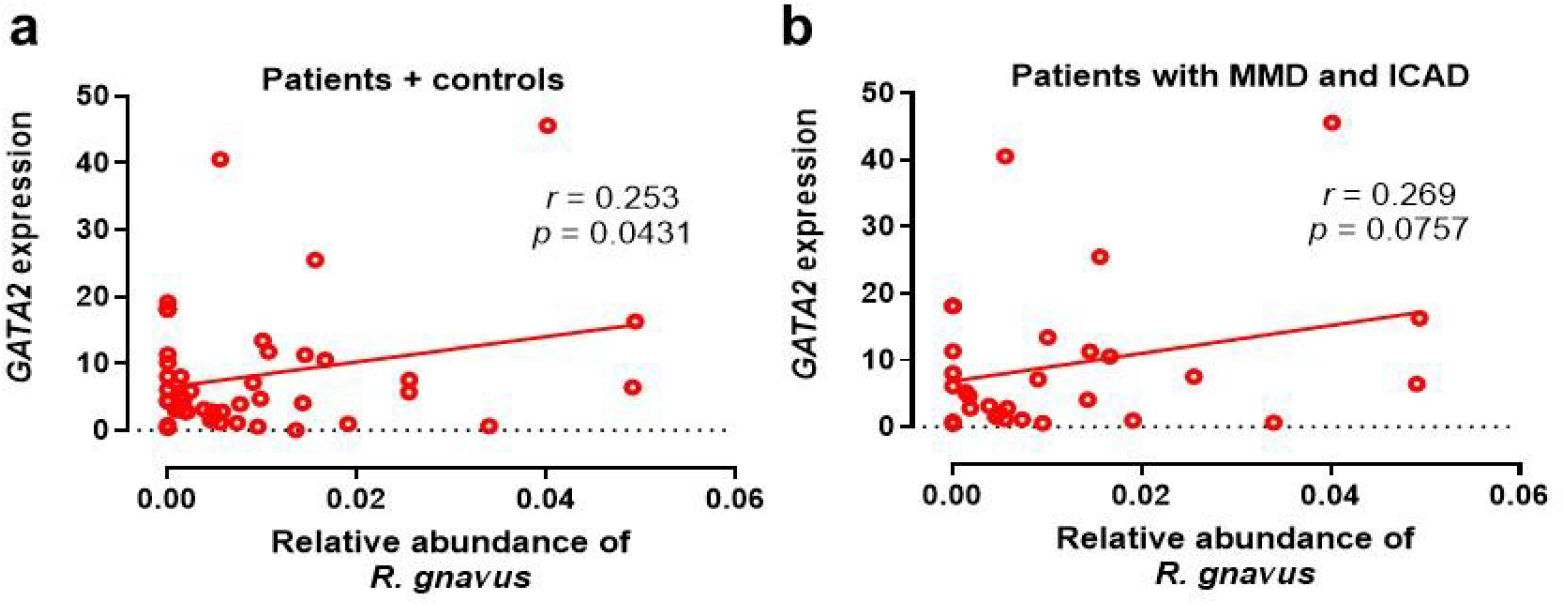
Peripheral Blood *GATA2* Expression Showed a Positive Correlation with the Relative Abundance of *Ruminococcus gnavus* in the Gut Microbiota. (a) *GATA2* expression displayed a linear correlation with the relative abundance of *Ruminococcus gnavus*. (b) This trend persisted in the case-only analysis, although it was not statistically significant.

## DISCUSSION

By comparing the transcriptome profiles of patients with moyamoya disease (MMD) and unaffected mutation carriers in highly aggregated familial MMD families, we identified differentially expressed gene networks that could potentially influence the penetrance of the *RNF213* founder mutation. The networks identified through Bayesian network analysis included: (1) the lipid-leukocyte module, which regulates lipid metabolism and the development of leukocyte including eosinophils and mast cells (*GATA2*, *SLC45A3*, *HDC*, and *AKAP12*), (2) mitochondrial ribosomal proteins and apoptosis (*RPL23*, *RPS3A*, *RPL26*, and others), (3) cytotoxic T-cell immunity (*GZMA*, *APOBEC3H*, and *TRGC1*), and (4) EGFR signaling (*UBTD1*). Of these networks, we focused on the lipid-leukocyte module because it showed the strongest signal in Bayesian network analysis and was replicated in WGCNA analysis. We confirmed that the genes within the lipid-leukocyte module were tightly linked, with *GATA2* and *SLC45A3* showing a strong correlation. Among individuals with the p.R4810K mutation, *GATA2* expression was significantly higher in MMD patients compared to age- and sex-matched unaffected carriers. ROC analysis indicated that a *GATA2* expression level above 8.06 could distinguish MMD patients from unaffected carriers. In familial MMD with the p.R4810K mutation, all individuals with *GATA2* expression levels above 8.06 developed MMD, while only one individual with low *GATA2* expression did so. Notably, higher *GATA2* expression was significantly associated with an earlier age at onset. This observation is unlikely to be confounded by the age at which blood was collected, as *GATA2* expression showed positive correlations with age in control individuals. Higher *GATA2* expression was also associated with a greater proportion of bilateral disease distribution and a higher proportion of symptomatic onset, although significant linear correlation was not confirmed. Interestingly, *GATA2* expression was higher in patients with MMD compared to those with ICAD, and the association between *GATA2* and clinical features was more apparent when MMD and ICAD were analyzed together, suggesting that RNF213-related vasculopathy represents a disease spectrum. It is believed that factors beyond RNF213, including GATA2, contribute to variations in phenotype across this spectrum.

*GATA2* is a transcription factor that regulates the maintenance and expansion of hematopoietic stem cells and progenitor cells,^38–40^ and is essential for mast cell and megakaryocyte development ^41,42^. It is also expressed on endothelial cells and hemogenic endothelium during the developmental stage, playing a role in endothelial-to-hematopoietic transition and endothelial-to-mesenchymal transition.^43^ A recent study showed that *GATA2* is a member of the lipid-leukocyte module, along with *SLC45A3*, *HDC*, *AKAP12*, *CPA3*, and *MS4A2*.^44^ Indeed, we confirmed that *GATA2* expression exhibited a significant linear correlation with *SLC45A3*. The lipid-leukocyte module is positively associated with triglycerides and negatively associated with high-density lipoprotein (HDL). This aligns with the finding that low HDL is a risk factor for MMD. Interestingly, palmitate levels are significantly increased in adipocytes through exogenous overexpression of *SLC45A3* in a dose-dependent manner,^45^ and *RNF213* is involved in regulating the lipotoxic effects of palmitate.^46^

*HDC* encodes the enzyme responsible for converting histidine into histamine, a well-known pro-inflammatory molecule secreted by basophils and mast cells.^47^ The STAT5-GATA2 pathway is crucial for basophil and mast cell differentiation and maintenance. *GATA2* activates the expression of *MS4A2*, a gene encoding the high-affinity IgE receptor, FcεRIβ, which is expressed on mast cells and basophils.^48^ These cells recognize antigens to secrete IL-6 and IL-13.^49^ IL-13 is highly expressed in extracranial blood vessels in MMD patients and negatively correlates with preoperative cerebral perfusion status.^50^ In an autopsy case of a patient who died from MMD, electron microscopy detected basophils and neutrophils, but not lipid-laden macrophages, in the endothelial layer at the stenotic lesions of the internal carotid arteries.^51,52^ Recent transcriptome analysis of the intracranial arteries of MMD patients suggested an upregulation of eosinophils and natural killer (NK) cells.^15^ These lines of evidence suggest that *GATA2* and its associated gene networks play a role in the pathophysiology of MMD.

The p.R4810K mutation in the *RNF213* gene is known to increase susceptibility not only to intracranial arterial disease but also to coronary artery disease (CAD) and pulmonary artery hypertension (PAH). Intriguingly, *GATA2* polymorphisms have been associated with familial early-onset coronary artery disease.^53^ *GATA2* regulates the CAD susceptibility gene *ADTRP* rs6903956 through preferential interaction with the G allele.^54^ *SLC45A3* has been reported to be associated with PAH, with PAH patients exhibiting higher mast cell infiltration compared to healthy controls and lower infiltration of naive CD4 T cells.^55^

We previously reported that a high abundance of *R. gnavus* is associated with MMD.^23^ Additionally, we observed that patients with MMD have a lower frequency of human herpesvirus 6 (HHV6) infection.^56^ Interestingly, *R. gnavus* has been shown to stimulate T cells to secrete IL-5 and IL-13,^37^ whereas HHV6 suppresses the levels of these cytokines.^57^ Given GATA2’s role in type 2 inflammation, including the development of eosinophils, basophils, and mast cells,^41,58^ we were interested in analyzing the relationship between *GATA2* expression and the relative abundance of *R. gnavus* in the gut. A moderate positive correlation was found among patients with MMD and ICAD. Although *R. gnavus* is associated with MMD and ICAD, there was no observed correlation with the penetrance of the p.R4810K mutation, suggesting that *R. gnavus* and *GATA2* are not in a direct one-to-one relationship but could be involved in disease progression through multiple distinct pathways.

Type 2 inflammation is particularly intriguing when considering the age of onset and gender differences in moyamoya disease. Androgen receptor signaling suppresses the expression of *GATA2* and *IL1RL1* (the IL-13 receptor).^59^ Interestingly, the age-of-onset patterns for moyamoya disease resemble those of asthma.^60^ Both exhibit a biphasic pattern, with the gender ratio shifting toward females in adulthood compared to childhood.^61^ In our study population, there was a trend of higher *GATA2* expression in boys during childhood and higher expression in women during adulthood, although this observation could be subject to selection bias.

Our study indicates that reduced mitochondrial ribosomal proteins may be associated with MMD. *RPS3A* positively regulates the mitochondrial function of human periaortic adipose tissue and is linked to coronary artery diseases.^62^ Notably, palmitate, whose levels are increased by *SLC45A3*, induces mitochondrial dysfunction, and cells expressing *GATA2* show reduced mitochondrial cristae density when observed under an electron microscope.^63^ This suggests that the lipid-leukocyte module and mitochondrial dysfunction could be related, as indicated by our Bayesian network analysis. In line with these observations, several reports have connected MMD with mitochondrial dysfunction. Moyamoya arteriopathy was observed in a child with impaired NADH-CoQ reductase activity,^64^ and in a patient with mitochondrial encephalopathy.^65^ Mitochondrial destruction has also been noted in circulating endothelial colony-forming cells from MMD patients.^66^ Regarding *RNF213*, the loss of mitochondrial matrix factors increases the expression of *RNF213*,^67^ and *RNF213* plays a role in non-mitochondrial oxygen consumption under the control of PTP1B.^68^

*GZMA* and *TRGC1* are expressed in cytotoxic T cells, effector memory T cells, and natural killer (NK) cells.^69^ *GZMA* is crucial for immune tolerance, and its deficiency leads to autoimmune conditions. *TRGC1* encodes the T cell receptor gamma, and reduced copy number has been associated with type 2 inflammation, including asthma.^70^ It has been reported that peripheral blood of MMD patients shows significantly increased levels of naive B cells and naive CD4 cells, along with a notable decrease in resting NK cells.^71^ *UBTD1* regulates cellular senescence through a UBTD1-Mdm2/p53 positive feedback loop,^72^ and is also known as a gene related to asthma exacerbation.^73^ Thus, not only the lipid-leukocyte module but also the effector T cell module and the *UBTD1* module are associated with type 2 inflammation.

We also found that *IL-1*β and *SOCS3* are connected in the network and that both are elevated. IL-1β has previously been reported to be elevated in the blood of patients with MMD. Ge et al. analyzed peripheral blood mononuclear cells (PBMCs) from MMD patients using single cell RNA sequencing and CyTOF, reporting that CD4+ effector memory T cells displayed features reminiscent of type 2 helper T (Th2) cells, characterized by elevated expression of GATA3, while GZMA+ CD8+ effector memory T cells were reduced.^12^ SOCS3 is known to promote the differentiation into GATA3+ Th2 cells,^74^ aligning with the result.

Additionally, since SOCS3 inhibits effector memory T cells that express GZMA,^74^ the connection between increased SOCS3 and decreased GZMA appears plausible. Ge and colleagues also reported an increase in STAT3 phosphorylation. IL-1β acts cooperatively with IL-6 to subvert the Foxp3-inducible Treg cell program in favor of Th17 cell development by phosphorylating STAT3 in the Foxp3 locus,^75^ providing a potential explanation for the increase in Th17 cells in patients with MMD. The IL-6/STAT3 signaling pathway is thought to play a role in transmitting inflammatory signals throughout the body, either through the bloodstream or the autonomic nervous system, suggesting that information related to inflammation, such as from the microbiome, could be transmitted systemically. On the other hand, SOCS3 can suppress STAT3, leading to a reduction in Th17 differentiation. Because IL-1β can either activate or inhibit SOCS3 depending on the context, the balance between Th2 and Th17 cells may vary with the situation. This implies that further research is needed to understand not only type 2 inflammation but also the regulation of the immunological microenvironment, including Th17 cells, in RNF213-related vasculopathy.

There are several limitations to this study. First, the study population is relatively small, indicating a need for further replication studies with larger sample sizes. Second, we analyzed peripheral blood mononuclear cells but not the blood vessel walls where the stenosis occurs.

Although it is challenging to obtain specimens from the vessel walls at the terminal portion of the internal carotid artery except in autopsy cases, vessel walls from peripheral middle cerebral arteries could be analyzed in future studies. Nonetheless, it is significant that our study demonstrated that peripheral blood can serve as a biomarker and that systemic circulation may be involved in the pathogenesis of MMD and the broader moyamoya spectrum, including RNF213-related vasculopathy. Third, this study did not directly establish a causal relationship between peripheral gene expression and MMD. Functional studies are needed to assess whether overexpression of *GATA2* induces MMD phenotypes. Finally, we did not test whether high *GATA2* expression is associated with an increased incidence of MMD among individuals with wild-type *RNF213*. Given that many family members with wild-type *RNF213* who exhibited high *GATA2* expression did not develop MMD, the impact of *GATA2* expression may be less evident in patients with wild-type *RNF213*. Further research is needed to explore these aspects.

### Conclusions

Our data suggest that higher *GATA2* expression is associated not only with increased penetrance of the p.R4810K mutation but also with a more severe form of MMD, characterized by symptomatic onset, an earlier age at onset, and a higher likelihood of bilateral involvement. The functionality of the gene network points to type 2 inflammation and mitochondrial dysfunction potentially playing a role in the pathogenesis of MMD spectrum and RNF213-related vasculopathy.

## Data Availability

All data produced in the present study are available upon reasonable request to the authors

## Author contributions

Conception and design: YM. Acquisition of data: YM, TK, Y.Oichi, T.Funaki. Analysis: TK, II. Analysis and interpretation of data: YM, MT, NS, YT. Drafting the article: YM, YT, NS, TK. Critically revising the article: TF, AK, Y.Okuno, SM, YA. Reviewed submitted version of manuscript: all authors. Statistical analysis: YM, MT, NS. Administrative/technical/material support: YM. Study supervision: AK, Y.Okuno, SM, YA.

## Acknowledgment

We are grateful to all the participants who donated their blood samples for this study.

AK holds a patent for RNF213 (JPWO2011049207A1), “Moyamoya disease-related genes and their use”. We filed a patent application for a method of predicting the onset of moyamoya disease by measuring GATA2 levels (YM, AK, O.Yasushi, SM). The other authors report no conflicts of interest.

## Availability of data and material

All data produced in the present study are available upon reasonable request to the authors.

## Abbreviations

MMD: moyamoya disease
ICAD: (non-moyamoya) intracranial large artery disease
LPS: lipopolysaccharide
R. gnavus: Ruminococcus gnavus
BN: Bayesian network
NNSR: Neighbor Node Sampling and Repeat
ECv: Edge Contributing value
WGCNA: Weighted Gene Co-Expression Network Analysis
SFTI: scale-free topology fit index
ANOVA: Analysis of Variance
ROC: Receiver operating characteristic
DEGs: Differentially expressed genes
AUC: area under the curve
CAD: Coronary artery disease
PAH: Pulmonary artery hypertension
HDL: high-density lipoprotein
PBMCs: Peripheral blood mononuclear cells
Th2: type 2 helper T

