## Supplemental files for "Peripheral blood *GATA2* expression impacts *RNF213* mutation penetrance and clinical severity in moyamoya disease"

**Supplemental Materials**

**Supplementary Table 1. Primers for RT-qPCR**

| **Gene** | **Forward** | **Reverse** |
| --- | --- | --- |
| *GATA2* | CTGCACATGGGTCATTGGTG | CGGAACCGGAAGATGTCCAA |
| *SLC45A3* | CCCCCAAAATGCCTAACCCA | GTGGTTAGGGAAGCCGTTGA |
| *RPS18* | CCTTTGCCATCACTGCCATT | TGATCACACGTTCCACCTCA |

**Supplementary Figure 1.**

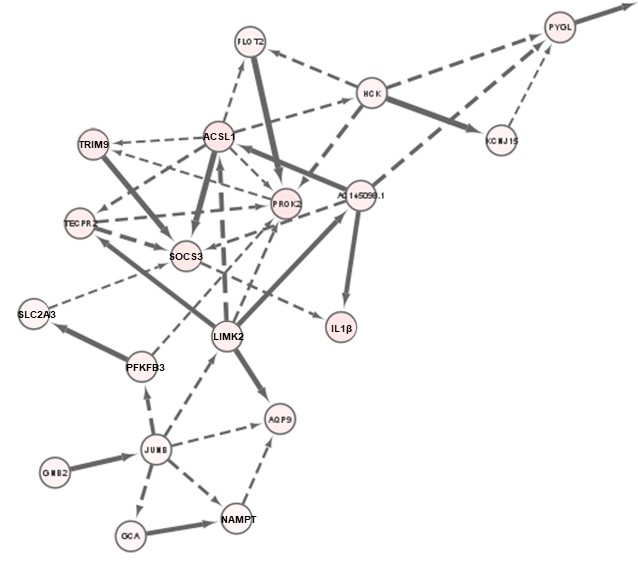

Among the molecular networks related to the penetrance of the R4810K mutation shown in Figure 2, we extracted the subnetwork indicated by the dotted circle. Although it does not meet the threshold of ΔECv >1.0, it forms an interesting network that includes IL-1β and SOCS3.

**Supplementary Figure 2.**

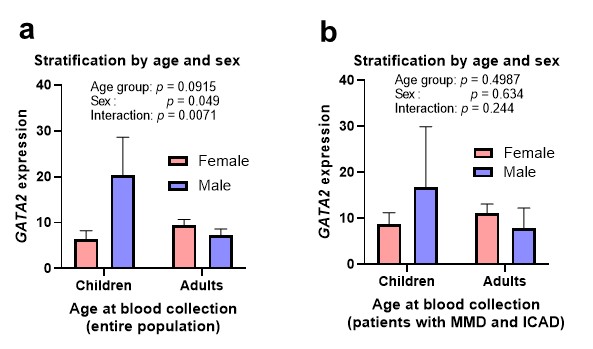

When stratifying *GATA2* expression by gender and age (children and adults), expression levels were higher in boys during childhood and slightly higher in women during adulthood, with a significant interaction effect. In an analysis limited to patients, the same trend was observed, although the difference was not statistically significant.

**Supplementary Figure 3.**

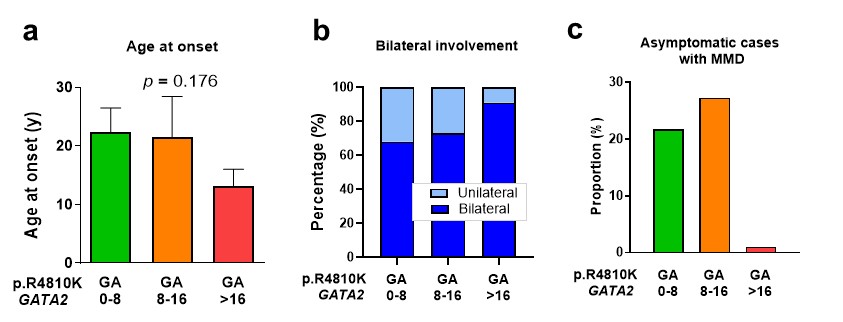

In Figure 4, the analysis was conducted on RNF213-related vasculopathy, including MMD and ICAD. Here, we present the results of an analysis limited to MMD.
